# Effect of Neoadjuvant Chemotherapy on Breast Conservative Surgery of Breast Cancer

**DOI:** 10.1101/2024.10.07.24314991

**Authors:** Mohamed Mostafa M Ahmed, Kamal Abdelrahman Abosenna, Basma Ahmed Mohamed, Noha Mohamed Nagla

## Abstract

**Background:** Breast conserving surgery (BCS) has been established as a safe and effective alternative way of total (modified radical) mastectomy for achieving loco regional control of female invasive breast cancer. There are still obstacles on generalizing neoadjuvant chemotherapy as a primary treatment of breast cancer even on early tumors, including large presenting tumors size relative to breast size especially on small breast sizes. Introducing preoperative (neoadjuvant) chemotherapy (NACT) has been argued to increase rates of neoadjuvant chemotherapy because of decreasing the overall tumor’s size.

**Aim of this study:** The aim of this work is to evaluate the efficacy of neoadjuvant chemotherapy on rates of breast conserving surgery on different molecular subgroups of breast cancer.

**Patients and methods:** Record of total of 44 patients with pathologically proven invasive breast cancer, who received neoadjuvant chemotherapy were included in this study. Eligibility of patients for breast conserving surgery before and after receiving neoadjuvant chemotherapy was measured in different molecular subtypes of breast cancer.

**Results:** Neoadjuvant chemotherapy increased eligibility for breast conserving surgery from 29% pre neoadjuvant chemotherapy to 77% post neoadjuvant chemotherapy. Highest rates of eligibility to neoadjuvant chemotherapy post neoadjuvant chemotherapy were achieved in HER-2 positive and Luminal A patients. Downstaging of the whole TNM disease stage decreased secondary to neoadjuvant chemotherapy in 89% of total study population. Luminal B patients showed the highest rates of downstaging. The average decrease in tumor size was 65.8% in response to neoadjuvant chemotherapy. HER-2 positive and triple negative patients showed the highest complete pathological response rates at 60.0% and 54.5% (p = 0.008).

**Conclusion:** Neoadjuvant chemotherapy increased the rate of breast conserving surgery with variation in response depending on the molecular subtype of the tumor. Her2+ tumors were the most sensitive subtypes to neoadjuvant chemotherapy with the highest breast conserving surgery eligibility following neoadjuvant chemotherapy.

## 1- Introduction

Worldwide, breast cancer is the most incident cancer among women with an estimated 2.3 million annual new cases, about 11.7% of all cancer cases. It is the most incident between Egyptian women and the second most common in both genders following liver cancer **(1,2)**. Breast-conserving surgery (BCS) followed by whole breast radiotherapy (WBRT) is a recognized treatment approach for early-stage breast cancer, offering comparable overall survival outcomes to mastectomy in a diverse age range of patients **(3)**.

Neoadjuvant (preoperative) chemotherapy (NACT) presents numerous benefits compared to the traditional sequence of surgery followed by adjuvant chemotherapy. Substantial evidence indicates that the implementation of neoadjuvant systemic chemotherapy has led to enhancements in overall survival (OS) rates and disease-free survival (DFS) of breast cancer patients. Further research is required to fully understand the impact of NACT on surgical decisions and BCS utilization in different breast cancer’s molecular subtypes **(4)**.

## 2- Patients and methods

Records of patients who received NACT at Al-Zahraa university hospital and Dar-Alsalam cancer hospital both based on Cairo, Egypt in the period between January 2023 and December 2023 were reviewed. Patients with absolute contraindication to BCS were excluded. Electronic medical records were chronologically followed up from the presenting examination sheet, imaging and pathology records, multi-disciplinary team (MDT) decision before starting NACT, follow up imaging records, MDT decision following completion of the planned NACT regimen, and post-surgery histopathology specimen reports data were collected.

### 2.1. Inclusion criteria

Female patients with invasive breast cancer, who received preoperative systemic therapy (Neoadjuvant Chemotherapy) were included to the study.

### 2.2. Exclusion criteria

Pregnant patients, patients with multi-centric breast cancer, patients with inflammatory breast cancer, and patient with proven metastasis before or after receiving NACT were excluded from the study.

### 2.3. Ethical consideration

The study involved data from the patients’ records regarding disease progression and treatments given. All patients’ data was confidential and no patient’s names or personal identifiers were disclosed to anybody. The study was be approved by the research ethics committees of general surgery department at Al-Zahraa university hospital, and of Faculty of medicine for girls, Al-Azhar university, Cairo, Egypt.

### 2.3. Patients’ subgroups

Forty-four patients were classified based on the molecular classification of breast cancer. Molecular subgroups included 10 patients with luminal A tumors, 13 with luminal B, 10 with HER2+, and 11 with triple negative (basal-like), comprising the total study population. Each molecular subgroup was classified according to the following criteria:

#### 1- Luminal A subgroup

ER positive, PR positive, HER-2 negative, and Ki67 proliferative index low.

#### 2- Luminal B subgroup

a. HER-2 negative: ER positive, PR negative (or low), HER-2 negative, and Ki67 proliferative index high.
b. HER-2 positive: ER positive, PR positive (or negative), HER-2 enriched, and Ki67 proliferative index varies.

#### 3- HER-2 positive subgroup

ER negative, PR negative, and HER-2 enriched.

#### 4- Triple negative breast cancer (TNBC) / (basal-like)

ER negative, PR negative, and HER-2 negative.

### 2.4. Measures

Each patient’s medical record was reviewed to gather clinical measures, including demographic characteristics, the maximum radiological tumor diameter at presentation, and the TNM stage. Immunohistochemical receptor status was assessed, focusing on estrogen receptor (ER), progesterone receptor (PR), HER2, and Ki67, alongside the classification of each case into molecular subtypes. Additionally, the tumor’s pathological type and grade were documented. The surgical decisions for each patient, both before and after receiving neoadjuvant chemotherapy (NACT), were also recorded to evaluate treatment outcomes.

The treatment outcome measures assessed were the number of breast-conserving surgeries performed after receiving neoadjuvant chemotherapy (NACT), the rate of pathological complete response post-NACT, the radiological change in tumor size from the initial presentation to post-NACT, and the rate of TNM stage downgrading after NACT.

### 2.5. Multi-disciplinary team

Surgical decisions were made by a multidisciplinary team comprising a surgery consultant, medical oncology consultant, radiation oncology consultant, pathology consultant, radiology consultant, and a clinical nutritionist.

### 2.6. Statistical Analysis

Data were coded and entered using the statistical package for the Social Sciences (SPSS) version 28 (IBM Corp., Armonk, NY, USA). Data was summarized using mean, standard deviation, median, minimum, and maximum in quantitative data and using frequency (count) and relative frequency (percentage) for categorical data. Comparisons between quantitative variables were done using the non-parametric Kruskal-Wallis and Mann-Whitney tests. For comparing categorical data, Chi square (χ2) test was performed. Exact test was used instead when the expected frequency is less than 5. P-values less than 0.05 were considered to be statistically significant.

## 3- Results

A total of 44 patients were included, with a mean age of 49.68 ± 11.16 years and a mean BMI of 29.61 ± 4.27. Of the patients, 52.3% were post-menopausal, 31.8% had a positive family history, 31.8% had diabetes, and 22.7% had hypertension. Left breast cancer was found in 52.3% of patients, while 6.8% had bilateral tumors. The most affected quadrant was the upper outer (52.3%), followed by the upper inner (20.5%), central (13.6%), lower inner (9.1%), and lower outer (4.5%). Unilateral breast cancer was present in 93.2% of patients **(Table 1)**.

**Table 1:** Main demographic data of the total study population.

| Variables | Mean ± SD (Min. – Max.) | |
| --- | --- | --- |
| Age (years) | $49.68 \pm 11.16$ (29 – 77) | |
| Body Mass Index (BMI) | $29.61 \pm 4.27$ (22 – 39) | |
| Variables | Count (total n=44) | Percent (%) |
| **Menopause** |  |  |
| Premenopausal | 21 | 47.7% |
| Postmenopausal | 23 | 52.3% |
| **Family History** |  |  |
| Positive | 14 | 31.8% |
| Negative | 30 | 68.2% |
| **Comorbidities** |  |  |
| Diabetes Mellitus | 14 | 31.8% |
| Hypertension | 10 | 22.7% |
| **Affected Breast Side** |  |  |
| Right breast | 21 | 47.7% |
| Left breast | 23 | 52.3% |
| **Affected Breast Quadrant** |  |  |
| Upper Outer Quadrant | 23 | 52.3% |
| Upper Inner Quadrant | 9 | 20.5% |
| Lower Outer Quadrant | 2 | 4.5% |
| Lower Inner Quadrant | 4 | 9.1% |
| Central Quadrant | 6 | 13.6% |
| **Laterality** |  |  |
| Unilateral | 41 | 93.2% |
| Bilateral | 3 | 6.8% |

The majority of the patients presented with advanced-stage disease. Tumor classification showed that 47.7% were T2, 18.2% were T3, and 29.5% were T4. Regarding lymph node involvement, 50% of patients were N1, 20.5% were N0, and 9.1% were N3, with all patients classified as M0. Overall, 27.3% were in stage IIIA, 25% in stage IIB, and 25% in stage IIIB. Pathologically, 90.9% of cases were invasive ductal carcinoma (IDC), with grade II tumors in 88.6% and grade III in 11.4%. Paget’s disease of the nipple was found in 6.8% of patients **(Table 2)**.

**Table 2:** Combined Disease Stage, TNM Classification, and Pathological Assessment of Study Population.

| Variable | Count<br>(total = 44) | % |
| --- | --- | --- |
| **Tumor** |  |  |
| T0 | 0 | 0.0% |
| T1 | 2 | 4.5% |
| T2 | 21 | 47.7% |
| T3 | 8 | 18.2% |
| T4 | 13 | 29.5% |
| **Nodes** |  |  |
| N0 | 9 | 20.5% |
| N1 | 22 | 50.0% |
| N2 | 9 | 20.5% |
| N3 | 4 | 9.1% |
| **Metastasis** |  |  |
| M0 | 44 | 100.0% |
| **Overall TNM Stage** |  |  |
| Stage I | 1 | 2.3% |
| Stage IIA | 5 | 11.4% |
| Stage IIB | 11 | 25.0% |
| Stage IIIA | 12 | 27.3% |
| Stage IIIB | 11 | 25.0% |
| Stage IIIC | 4 | 9.1% |
| **Pathological Type** |  |  |
| IDC * | 40 | 90.9% |
| ILC * | 2 | 4.5% |
| IMC * | 1 | 2.3% |
| IMPC * | 1 | 2.3% |
| **Pathological Grade** |  |  |
| Grade II | 39 | 88.6% |
| Grade III | 5 | 11.4% |
| **Paget's Disease of the Nipple** | 3 | 6.8% |

\* IDC=invasive ductal carcinoma, ILC=invasive lobular carcinoma, IMC=invasive mucinous carcinoma, IMPC=invasive micropapillary carcinoma

When comparing presenting disease stage between sub-groups, notable differences emerged. In Luminal A, most patients were in early stages (I to IIB), with 90% having minimal lymph node involvement (N0-N1). In Luminal B, patients were more evenly distributed across stages, including advanced stages (IIIA and IIIC), with 62% showing significant lymph node involvement (N1-N2), and all were M0. The HER2+ subgroup had patients mainly in stages IIA to IIIB, with 100% having significant lymph node involvement (N1) and no metastasis (M0). TNBC patients were distributed across stages IIA to IIIC, with a tendency towards advanced stages (IIIB), 73% having significant lymph node involvement (N2-N3), and all were M0. Paget’s Disease was rare overall, absent in Luminal A but present in 7.7% of Luminal B, 10.0% of HER2+, and 9.1% of TNBC patients. This indicates diverse breast cancer characteristics across molecular subtypes **(Table 3)**.

**Table 3:** Distribution of Breast Cancer Pathological Type and Disease Stage Across Molecular Subtypes.

| Variable | Luminal A<br>N= 10 | Luminal B<br>N= 13 | Her2+<br>N= 10 | Triple<br>Negative<br>N= 11 | P-value |
| --- | --- | --- | --- | --- | --- |
| Pathological Type |  |  |  |  |  |
| IDC * | 6 | 13 | 10 | 11 | 0.003** |
| ILC * | 2 | 0 | 0 | 0 |  |
| IMC * | 1 | 0 | 0 | 0 |  |
| IMPC * | 1 | 0 | 0 | 0 |  |
| Stage |  |  |  |  |  |
| Stage I | 1 | 0 | 0 | 0 | 0.914 |
| Stage IIA | 1 | 1 | 1 | 2 |  |
| Stage IIB | 4 | 2 | 3 | 2 |  |
| Stage IIIA | 2 | 5 | 3 | 2 |  |
| Stage IIIB | 2 | 3 | 3 | 3 |  |
| Stage IIIC | 0 | 2 | 0 | 2 |  |
| Tumor |  |  |  |  |  |
| T1 | 1 | 0 | 1 | 0 | 0.470 |
| T2 | 4 | 7 | 3 | 7 |  |
| T3 | 3 | 2 | 3 | 0 |  |
| T4 | 2 | 4 | 3 | 4 |  |
| Nodes |  |  |  |  |  |
| N0 | 5 | 1 | 0 | 3 | 0.003** |
| N1 | 4 | 6 | 10 | 2 |  |
| N2 | 1 | 4 | 0 | 4 |  |
| N3 | 0 | 2 | 0 | 2 |  |
| Metastasis |  |  |  |  |  |
| M0 | 10 | 13 | 10 | 11 | ----- |
| Paget's Disease |  |  |  |  |  |
|  | 0 | 1 | 1 | 1 | 1 |

\*IDC=invasive ductal carcinoma, ILC=invasive lobular carcinoma, IMC=invasive mucinous carcinoma, IMPC=invasive micropapillary carcinoma, \*\*Statistically p-value at below 0.05

**Figure 1** compares BCS eligibility before and after neoadjuvant chemotherapy (NACT) across molecular subgroups and the total population. Before NACT, eligibility was 30.0% in Luminal A, 30.8% in Luminal B, 20.0% in HER2+, and 36.4% in Triple Negative, with an overall rate of 29.5%. After NACT, eligibility improved to 80.0% in Luminal A, 76.9% in Luminal B, 90.0% in HER2+, and 63.6% in Triple Negative, with a total rate of 77.3%. This indicates a significant overall increase in BCS eligibility post-NACT, though subgroup differences were not statistically significant (p=0.614).

**Figure 1:**
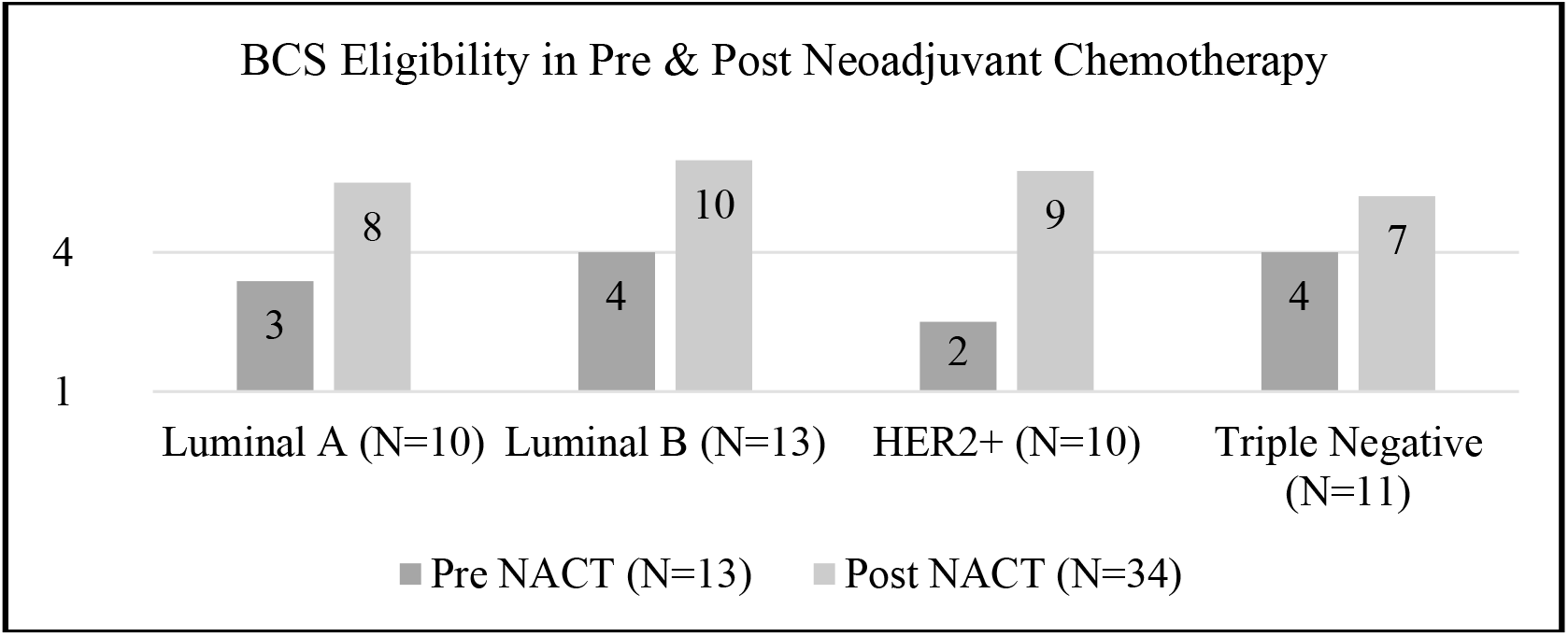
Comparison of breast conserving surgery eligibility before and after neoadjuvant chemotherapy between study molecular subgroups, *p-value= 0.614*

**Table 4** shows that surgical decisions varied across molecular subgroups. For breast-conserving surgery (BCS), quadrantectomy was performed in 25% of patients overall, with 20% in Luminal A, 15% in Luminal B, 20% in HER2+, and 46% in Triple Negative. Lumpectomy was done in 52% of patients overall, with higher rates in Luminal A (60%), Luminal B (62%), and HER2+ (70%). Wire-guided procedures were used in 52% of patients, applicable to either lumpectomy or quadrantectomy, with significant variation: Luminal A (50%), Luminal B (62%), HER2+ (80%), and Triple Negative (18%), showing a significant difference (p=0.034). Total mastectomy was performed in 32% of patients, with 20% in Luminal A, 23% in Luminal B, 10% in HER2+, and 36% in Triple Negative, with no significant differences among groups (p=0.614).

**Table 4:** Final Surgical Decisions Across Molecular Subgroups.

| Final Surgical Decision | Luminal A |  | Luminal B |  | HER2+ |  | Triple Negative |  | Total (N=44) |  | P-Value |
| --- | --- | --- | --- | --- | --- | --- | --- | --- | --- | --- | --- |
|  | No. | % | No. | % | No. | % | No. | % | No. | % |  |
| **Quadrantectomy** | 2 | 20% | 2 | 15% | 2 | 20% | 5 | 46% | 11 | 25 % | 0.351 |
| **Lumpectomy** | 6 | 60% | 8 | 62% | 7 | 70% | 2 | 18% | 23 | 52 % | 0.074 |
| **Total Mastectomy** | 2 | 20% | 3 | 23% | 1 | 10% | 4 | 36% | 10 | 32% | 0.614 |

The study demonstrated an average tumor size reduction of 65.88% post-neoadjuvant chemotherapy (NACT), with the highest reduction in the Triple Negative subgroup (77.97%), followed by HER2+ (66.85%), Luminal A (64.43%), and Luminal B (55.18%). The overall mean tumor size decreased from 4.25 cm to 1.45 cm, with a reduction of 2.79 cm. HER2+ tumors had the largest pre-NACT sizes, while Triple Negative tumors had the most significant post-NACT size reduction, highlighting their higher chemosensitivity, along with HER2+ tumors **(Table 5)**.

**Table 5:** Tumor Size Reduction Before and After Neoadjuvant Chemotherapy.

| Tumor Size (cm) | Total Study Population | Luminal A | Luminal B | HER2+ | Triple Negative | P-value |
| --- | --- | --- | --- | --- | --- | --- |
| **Pre-NACT** |  |  |  |  |  |  |
| Mean | 4.25 | 3.92 | 4.11 | 4.89 | 4.13 | 0.984 |
| Median | 3.55 | 3.55 | 3.70 | 3.55 | 3.40 |  |
| Range (Min - Max) | 1.20 - 10.00 | 1.30 - 7.30 | 2.30 - 9.60 | 1.20 - 10.00 | 1.50 - 8.60 |  |
| **Post-NACT** |  |  |  |  |  |  |
| Mean | 1.45 | 1.39 | 1.84 | 1.62 | 0.91 | 0.503 |
| Median | 0.85 | 1.50 | 1.80 | 0.00 | 0.00 |  |
| Range (Min - Max) | 0.00 - 8.50 | 0.00 - 3.50 | 0.00 - 7.50 | 0.00 - 8.50 | 0.00 - 3.90 |  |
| **Size Reduction** |  |  |  |  |  |  |
| Mean | 2.79 | 2.53 | 2.27 | 3.27 | 3.22 | 0.727 |
| Percentage | 65.88% | 64.43% | 55.18% | 66.85% | 77.97% |  |

BCS = Breast Conserving Surgery, NACT = Neoadjuvant Chemotherapy

When assessing tumor’s pathological response to neoadjuvant chemotherapy according to the molecular subtypes of the tumor, the HER2+ subgroup demonstrated the highest complete response rate at 60.0%, followed by the Triple Negative subgroup at 54.5%, which are thought to have more chemosensitivity than luminal tumors. In contrast, the Luminal A subgroup exhibited a complete response rate of 20.0%, while the Luminal B subgroup had the lowest rate at 7.7% **(Table 6)**. The statistical analysis revealed a significant difference in complete pathological response among the molecular subgroups (p = 0.008).

**Table 6:** Comparison of Pathological Response and Overall Disease TNM Downstaging in Response to Neoadjuvant Chemotherapy Between Study’s Molecular Subgroups.

| Variables | Molecular classification subgroups |  |  |  |  |  |  |  | *P*-value |
| --- | --- | --- | --- | --- | --- | --- | --- | --- | --- |
|  | Luminal A |  | Luminal B |  | HER2+ |  | Triple Negative |  |  |
|  | n. | % | n. | % | n. | % | n. | % |  |
| Pathological Response |  |  |  |  |  |  |  |  | 0.008* |
| Complete | 2 | 20.0% | 1 | 7.7% | 6 | 60.0% | 6 | 54.5% |  |
| Partial | 7 | 70.0% | 12 | 92.3% | 3 | 30.0% | 3 | 27.3% |  |
| Stationary | 1 | 10.0% | 0 | 0.0% | 1 | 10.0% | 1 | 9.1% |  |
| Progressive | 0 | 0.0% | 0 | 0.0% | 0 | 0.0% | 1 | 9.1% |  |
| TNM Down Staging |  |  |  |  |  |  |  |  | 0.855 |
| Yes | 9 | 90.0% | 12 | 92.3% | 8 | 80.0% | 10 | 90.9% |  |
| No | 1 | 10.0% | 1 | 7.7% | 2 | 20.0% | 1 | 9.1% |  |

\*Statistically *p*-value at below 0.05

In the study, 88.6% of patients experienced downstaging post-neoadjuvant chemotherapy (NACT), with the highest frequency in the Luminal B subgroup **(Table 6)**. Downstaging rates were high across all molecular subgroups: 90.0% in Luminal A, 92.3% in Luminal B, 80.0% in HER2+, and 90.9% in Triple Negative. The differences between subgroups were not statistically significant (p=0.855).

## 4- Discussion

Breast-conserving surgery (BCS) is a proven, safe, and effective alternative to total mastectomy for controlling localized breast cancer. Research has consistently shown that BCS provides comparable outcomes in terms of overall survival and disease-free survival **(5)**. However, challenges remain in widely adopting BCS as a primary treatment, even for some cases with early-stage tumors. These challenges include a large tumor size relative to breast size, especially in patients with smaller breasts, and advanced T stage at diagnosis, even in early TNM stages **(6)**.

Preoperative neoadjuvant chemotherapy (NACT) has been proposed as a solution to increase BCS rates by reducing tumor size before surgery. NACT offers the benefit of downstaging tumor size prior to surgery, enabling less invasive surgical procedures. Additionally, NACT helps in better intraoperative tumor identification and reduces the likelihood of significant residual disease **(7)**.

**Richter et al. (8)** reported a mean age of 50 years and a mean BMI of 28.08 (SD 5.59) among patients with breast cancer who received NACT. With respect to affected breast quadrant, **Mostafa et al. (9)** identified the upper outer quadrant as the most commonly affected site (53.3%) in breast cancer.

The findings of this study are consistent, with a mean age of 49.68 (SD 11.16) and a mean BMI of 29.61 (SD 4.27) in conceding with **Richter et al. (8)**. Similarly, this study found that the upper outer quadrant was the most frequently involved (52%), followed by the lower outer quadrant (20%) and the central quadrant (14%) in conceding with **Mostafa et al. (9)** regarding the most affected quadrant.

**O’Halloran et al. (10)**, in their review of the effect of NACT on surgical practice, patient outcomes, and patterns of disease recurrence in breast cancer, reported that the majority of patients receiving NACT (77.6%) had T2 to T3 stages. The most common overall TNM stage was stage II (50.4%), followed by stage III (46.4%). **Richter et al. (8)**, also reported that the majority of their patients presented with clinical stage III (67.3%).

In the present study, the most frequent T stage was T2 (47.7%), followed by T4 (29.5%), while the most common TNM stage was stage III (61.36%), followed by stage II (36.36%). This study population exhibited higher T stages and overall TNM stages compared to the results reported by **O’Halloran et al. (10)**. Additionally, the results of this study are conceding with those of **Richter et al. (8)**, who also reported that the majority of their patients presented with clinical stage III. This finding highlights the need for higher TNM stage patients to receive NACT.

Regarding the pathological type of breast cancer, **Zaher et al. (11)** reported invasive ductal carcinoma accounting for the vast majority of cases (89.8%). However, the pathological grade differed, as their most encountered grade was grade III (49.7%), followed by grade II (38.8%),

In this study, the most common pathological type was invasive ductal carcinoma in 90.9%, and grade II was the most prevalent (88.6%), followed by grade III (11.4%). These results are conceding with results of **Zaher et al. (11)** in the pathological type but different in the grade.

BCS eligibility following NACT was examined by **Petruolo et al. (12)**, who evaluated the conversion rates from BCS ineligibility to eligibility in patients with large tumors. In their study, 69% were initially ineligible for BCS, while 31% were considered borderline candidates. Following NACT, 69% of the initially ineligible patients became eligible for BCS. Overall, 48% of patients who were initially deemed ineligible for BCS due to large tumor size were able to avoid mastectomy after NACT. **Asselain et al. (13)** also found that NACT increased BCS rates to 65%, compared to 49% in patients who underwent upfront surgery followed by adjuvant chemotherapy.

Results from this study reported that after NACT, BCS eligibility increased from 13 patients to 34 patients of whole study population. At the start of the study, 31 patients (70.5%) of the study population were non-eligible for BCS, 21 of them (47% of the total population) became eligible for BCS after NACT and avoided total mastectomy. NACT increased eligibility for BCS from 29.5% to 77.3% of the total study population in conceding with **Petruolo et al. (12)** and **Asselain et al. (13)**.

**Golshan et al. (14)**, in a prospective study of 634 patients with triple-negative breast cancer (TNBC), reported that neoadjuvant systemic therapy enabled breast-conserving surgery (BCS) in 53.2% of these women.

In this study, we observed a significant increase in BCS eligibility following neoadjuvant chemotherapy (NACT) across all molecular subgroups. Prior to treatment, BCS eligibility ranged from 20.0% in HER2+ patients to 36.4% in TNBC patients in conceding with **Golshan et al. (14)**. After NACT, eligibility improved markedly, reaching 80.0% in Luminal A, 76.9% in Luminal B, 90.0% in HER2+, and 63.6% in TNBC. These findings underscore the positive impact of NACT in enhancing surgical options for patients across different molecular subgroups.

In the study by **Yamaguchi et al. (15)**, which evaluated neoadjuvant chemotherapy (NACT) in converting HER2-positive breast cancer patients from breast-conserving therapy (BCT) ineligible to BCS eligible, the average tumor size decreased from 3.27 ± 1.37 cm to 1.44 ± 1.19 cm. Following NACT, 49% of the population became eligible for BCS, with 48% undergoing BCS instead of total mastectomy. The proportion of patients deemed ineligible for BCS due to large tumor size dropped from 47% to 14%.

In this study’s results, NACT reduced tumor sizes from 4.25 ± 2.23 cm to 1.45 ± 1.87 cm, with an average size reduction of 2.79 ± 2.28 cm. This corresponds to a 65.88% reduction in tumor size across the total study population in conceding with **Yamaguchi et al. (15)**.

When studying size reduction on breast cancer subgroups, the HER2+ subgroup had larger initial tumor sizes compared to the other subgroups, while the triple-negative subgroup showed the lowest median post-NACT size, despite having the highest initial median size. Consequently, the triple-negative subgroup exhibited the greatest tumor size reduction, highlighting it, along with the HER2+ subgroup, as the most chemo-sensitive in our population in agreement with **Golshan et al. (14)** results.

Regarding tumor’s pathological response to NACT, **Schmitz et al. (16)** who reported higher complete response rates in HER2-positive (76.1%) and Triple Negative (56.4%) tumors compared to Luminal A (12.6%).

In this study, the tumor’s pathological response to NACT varied significantly across stages and molecular subtypes. The HER2+ subgroup had the highest complete pathological response (pCR) rate (60.0%), followed by Triple Negative (54.5%), while Luminal A and Luminal B had lower pCR rates (20.0% and 7.7%, respectively) in conceding with **Schmitz et al. (16)** results. This underscores the importance of molecular subtype in treatment planning.

Agreeing to the mentioned results, **Zaher et al. (11)** evaluated the impact of neoadjuvant systemic chemotherapy (NAC) on women with stage II to III breast cancer, reporting a pCR in 76.4% of patients. They observed higher pCR rates in HER2+ (77.4%) and Triple Negative Breast Cancer (61.8%) compared to Luminal A (28.6%) and Luminal B (46.2%). These findings are consistent with our results, particularly in the HER2+ and TNBC subgroups.

In this study, NACT greatly increased BCS eligibility across the population. Tumor sizes were significantly reduced, with the HER2+ and TNBC subtypes showing the highest responsiveness to chemotherapy. The TNBC subgroup, despite presenting with the largest tumor sizes, exhibited the greatest reduction in size, indicating its high chemo-sensitivity, similar to HER2+ patients. Pathological responses also varied by molecular subtype, with HER2+ and TNBC achieving the highest complete pathological response rates, while Luminal A and Luminal B subtypes showed more modest responses. These findings emphasize the critical role of molecular subtypes in predicting response to NACT and optimizing surgical treatment options **Loibl et al (3)**.

## 5- Conclusion

Administration of neoadjuvant chemotherapy has revolutionized the surgical management of female’s breast cancer. The most clearly established advantage of neoadjuvant chemotherapy is its ability to convert patients initially ineligible for breast conserving surgery into candidates for this treatment. This study provided insights into the efficacy of neoadjuvant chemotherapy in inducing downstaging of the disease across different study populations and molecular subgroups. Our preliminary results confirm that the neoadjuvant chemotherapy increases the chances of breast-conserving surgery in patients with early and advanced cancers. We believe that the key to successful breast conserving surgery after neoadjuvant chemotherapy are careful patient selection, and multidisciplinary coordination.

## Data Availability

All data produced in the present study are available upon reasonable request to the authors

